# Impact of COVID-19 restrictions on diabetes health checks and prescribing for people with type 2 diabetes: a UK-wide cohort study involving 618,161 people in primary care

**DOI:** 10.1101/2021.04.21.21255869

**Authors:** Matthew J. Carr, Alison K. Wright, Lalantha Leelarathna, Hood Thabit, Nicola Milne, Naresh Kanumilli, Darren M. Ashcroft, Martin K. Rutter

## Abstract

**OBJECTIVE:** To compare rates of performing NICE-recommended health checks and prescribing in people with type 2 diabetes (T2D), before and after the first COVID-19 peak in March 2020, and to assess whether trends varied by age, sex and deprivation.

**METHODS:** We constructed a cohort of 618,161 people with T2D followed between March and December 2020 from 1744 UK general practices registered with the Clinical Practice Research Datalink (CPRD; Aurum and GOLD databases). We focused on the following six health checks and prescribing: HbA1c, serum creatinine, cholesterol, urinary albumin excretion, blood pressure and BMI assessment, comparing trends using regression models and 10-year historical data.

**RESULTS:** In April 2020, in English practices, rates of performing health checks were reduced by 76-88% when compared to 10-year historical trends, with older people from deprived areas experiencing the greatest reductions. Between May and December 2020, the reduced rates recovered gradually but overall remained 28% and 47% lower compared to historical trends, with similar findings in other UK nations. In England, rates of prescribing of new medication fell during April with reductions varying from 10% (4-16%) for antiplatelet agents to 60% (58-62%) for antidiabetic medications. Overall, between March and December 2020, the overall rate of prescribing new diabetes medications was reduced by 19% (15-22%) and new antihypertensive medication by 22% (18-26%). Similar trends were observed in other UK nations, except for a reduction in new lipid-lowering therapy prescribing March to December 2020 (reduction: 16% (10-21%)).

**CONCLUSIONS:** Over the coming months, healthcare services will need to manage this backlog of testing and prescribing. Effective communications should ensure that patients remain engaged with diabetes services, monitoring and opportunities for prescribing, and make use of home monitoring and remote consultations.

## INTRODUCTION

The COVID-19 pandemic has had major health and economic effects across the world. So far in the UK, there have been more than 126,000 COVID-related deaths with disproportionate impacts in people with diabetes; in the early phase of the pandemic nearly a third of all COVID-related deaths occurred in people with diabetes.^1-3^

The impact on the NHS, and in particular on diabetes services, has been enormous, with the suspension of much routine care. As the COVID-19 pandemic continues, there is an urgent need to minimise the harm done through reduction of routine services and to prioritise care and resources to areas of greatest need.

In 2008, the National Institute for Health and Care Excellence (NICE) recommended nine essential ‘health checks’ or so-called ‘care processes’ that define high-quality diabetes care.^4^ NICE recommended that people with diabetes should have at least annual checks of weight, blood pressure, smoking status, HbA1c, cholesterol, creatinine, urinary albumin, retinopathy and feet. Since 2008, these health checks have been incorporated in National Diabetes Audits and have also been used very effectively in the Quality Outcomes Framework (QOF) to incentivise high-quality primary care services.

The management of type 2 diabetes occurs almost exclusively in primary care.^5^ Therefore, lower general practice attendance due to COVID-19 would likely restrict the ability to perform these essential health checks. Consequently, this could have adverse effects on patient safety and increase the risk of developing long-term diabetes-related complications.

There is limited data on the indirect consequences of the COVID-19 pandemic on diabetes health checks and prescribing in primary care. Therefore, we used a large primary care longitudinal dataset, broadly representative of the UK population, to compare the frequency of health checks and prescribing in people with type 2 diabetes, before and after the first nationwide COVID-19 lockdown in March 2020. We compared observed and predicted rates using data covering ten years prior to the pandemic.

Since older people and more socially disadvantaged groups have been disproportionally affected by COVID-19 infections, and since the same groups may be more adversely impacted by changes in healthcare delivery imposed by COVID-19, we aimed to study variation in outcomes by gender, age group and deprivation level.

## METHODS

### Data sources

We conducted a retrospective cohort study using primary care electronic health records obtained from the Clinical Practice Research Datalink (CPRD) *Aurum* and *GOLD* databases ^6,7^. The study population consisted of 965,964 patients with a diagnosis of type 2 diabetes; 824,698 patients from 1470 general practices in England, with a further 141,266 patients from 361 practices across Northern Ireland (16,408 patients in 40 practices), Scotland (69,935 patients in 208 practices), and Wales (54,923 patients in 113 practices). From the study population, 934,214 patients (from 1828 UK general practices) contributed to the estimation of the expected rates in the pre-COVID-19 period between January 2010 and February 2020.

The CPRD contains anonymised consultation records and includes patient demographic information, symptoms, diagnoses, medication prescriptions, and date of death. We also examined practice-level Index of Multiple Deprivation (IMD) quintiles,^6^ a measure representing an area’s relative level of deprivation, ranked within each UK nation.

### Definitions, measurements and clinical coding

To enable comparisons of rates before and after the start of the COVID-19 outbreak, we included patient records from January 2010 to establish long-term trends and patterns of seasonality. We focused primarily on reporting observed versus expected rates from 1^st^ March 2020 to 10^th^ December 2020. For the diabetes monitoring component of the study, we restricted our investigation to the following six care processes because there was a high level of confidence that that they had or had not been performed based on the available primary care records: HbA1c, serum creatinine, cholesterol, urinary albumin excretion, blood pressure, BMI measurement. It seemed possible that smoking status had been assessed and foot checks performed but not recorded in the practice records and therefore we did not report these outcomes. Eye screening for retinopathy is performed in the community and these data are not recorded routinely in the primary care record.

For the medication prescribing component of the study, we focused on medications commonly prescribed to patients with a diagnosis of type 2 diabetes: antidiabetics, antihypertensives, lipid-lowering drugs and antiplatelet agents. To compare prescribing behaviours before and after the start of the pandemic, we applied two distinct definitions: First, we assessed the prescribing of new medication within a three-year window of a first diagnosis of type 2 diabetes. Second, we assessed the overall prescribing rate (new and repeat) among patients with a prior diagnosis.

We also compared the incidence and event rates for eight separate strata by combining attributes of the study cohort via the dichotomisation of gender, age (less than 65 versus greater than or equal to 65), and IMD (quintiles 4 and 5 (most deprived) versus quintiles 1, 2, and 3) as shown in **Supplementary Table 1**.

Care processes were identified using Read/SNOMED/EMIS codes used in CPRD *GOLD* and *Aurum*. Medication prescribing events were identified using CPRD product codes linked to codes from the dictionary of medicines and devices (DM+D) (see https://clinicalcodes.rss.mhs.man.ac.uk).

In line with guidance from the CPRD’s central administration, data from the *Aurum* and *GOLD* databases were analysed separately, with data from *Aurum* restricted to English practices and *GOLD* providing information on practices in Northern Ireland, Scotland and Wales. The use of two discrete data sources also enabled independent replication of our findings. All code lists and medication lists were verified by two senior clinical academics (a diabetologist: MKR, and a senior academic pharmacist: DMA).

### Study design

For each patient, we defined a ‘period of eligibility’ for study inclusion which commenced on the latest of: the study start date (1st January 2010); the patient’s most recent registration with their practice; the date on which data from the practice was deemed to be ‘up-to-standard’ by the CPRD; the patient’s first diagnosis of type 2 diabetes. A patient’s period of eligibility ended on the earliest of: registration termination; the end of data collection from their practice; death. We also applied a ‘look-back’ period during which a patient was required to have been registered for at least a year prior to their diagnosis of type 2 diabetes. The denominator for the incidence and event rates was the aggregate person-months at risk for the whole eligible study cohort.

### Statistical Analysis

The data were structured in a time-series format with event counts and ‘person-months at risk’ aggregated (by year and month) with stratification by gender, age group, deprivation quintile and region (or nation in *GOLD*). Mean-dispersion negative binomial regression models were used to estimate expected monthly event counts from March 2020 onward based on antecedent trends since 2010. The natural logarithm of the denominator (person-months at risk) was used as an offset in each regression model. To account for possible seasonality and long-term linear trends, calendar month was fitted as a categorical variable and time as a continuous variable with the number of months since the start of the study serving as the unit of measurement. For each month studied, observed and expected event counts were converted to rates using the observed person-month denominator. The monthly expected rates, and their 95% confidence intervals, were plotted against the observed rates. As they share a common denominator, differences between expected and observed monthly rates are expressed as a percentage ‘rate reduction (or increase)’.

All data processing and statistical analyses were conducted using Stata version 16 (StataCorp LP, College Station, TX). We followed RECORD (REporting of studies Conducted using Observational Routinely-collected health Data) guidance ^10^.

## RESULTS

### Study cohort

Our focus was on the impact of the COVID-19 pandemic between March and December 2020. Using the inclusion criteria described in the Study Design, a mixed cohort was utilised consisting of those patients from the study population whose period of eligibility began before 1st March 2020 and those who became eligible for inclusion between 1st March 2020 and 10th December 2020. The study cohort was comprised of 618,161 people with type 2 diabetes from 1744 UK general practices. The median (IQR) age was 68 (58, 77) years, 44% were female and 25% lived in an area that was in the most deprived quintile compared to the rest of the UK.

#### Impacts of COVID-19 on care processes in T2D

In April 2020, in English primary care, the rate of performing health checks was reduced by 76-88% when compared to 10-year historical trends (**Figure 1)**, with similar reductions (74-88%) in other UK nations (**Supplementary Figure 1**).

**Figure 1.**
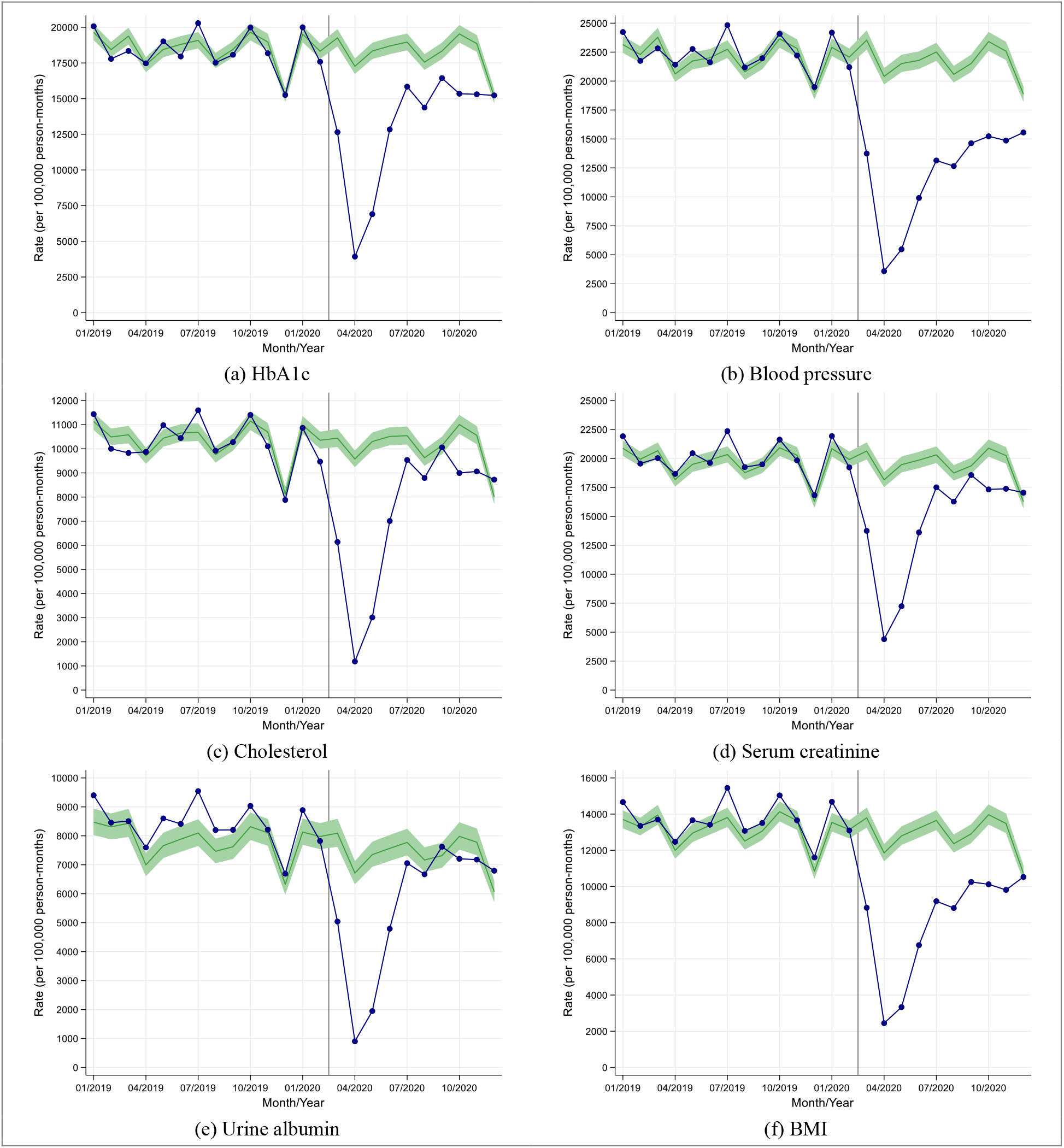
Observed and expected care process rates in people with type 2 diabetes during 2019 and 2020, in England

Although reductions in rates of performing health checks were similar by age, gender and socio-economic group, older people from deprived areas tended to have the greatest reductions in rates due to having higher background testing rates (England, **Figure 2;** other UK Nations, **Supplementary Figure 2**).

**Figure 2.**
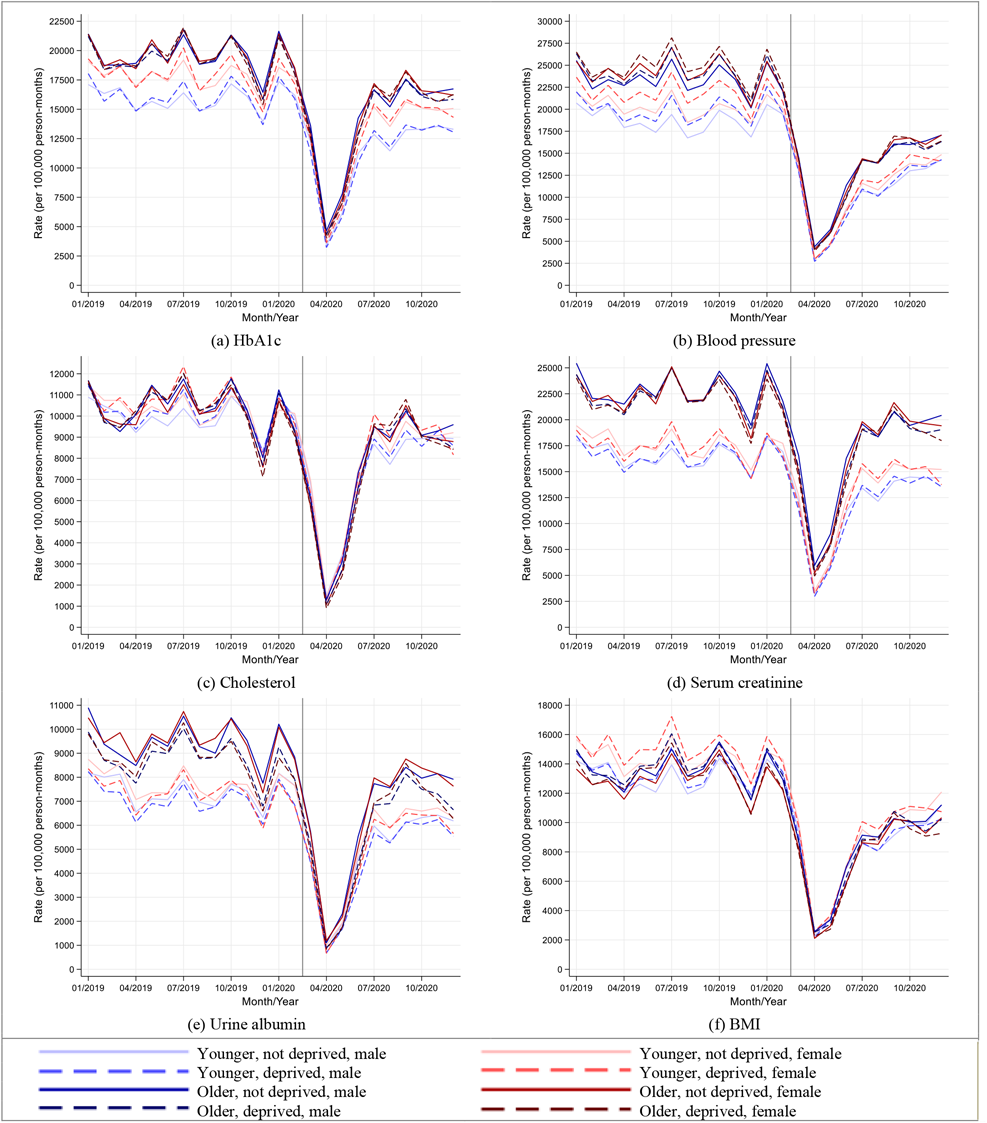
Stratified care process rates in people with type 2 diabetes during 2019 and 2020, in England

Between May and December 2020, the reduced rates of performing health checks recovered gradually though rates remained well below expected especially for blood pressure and BMI monitoring (England, **Figure 1;** other UK Nations, **Supplementary Figure 1**).

Overall in English practices, between 1^st^ March and 10^th^ December 2020, the rate of performing each of the health checks was reduced by between 28% and 47% compared to historical trends (**Table 1**); the most affected health check being blood pressure testing (rate reduction (95% CI): 47% (45-49%)) and the least affected being urine albumin monitoring (reduction: 28% (23-32%)). Similar trends were observed in other UK nations with rate reductions varying between 37-51% across the health checks (**Supplementary Table 2**); blood pressure monitoring being most significantly reduced (rate reduction: 51% (49-53%)).

**Table 1.**
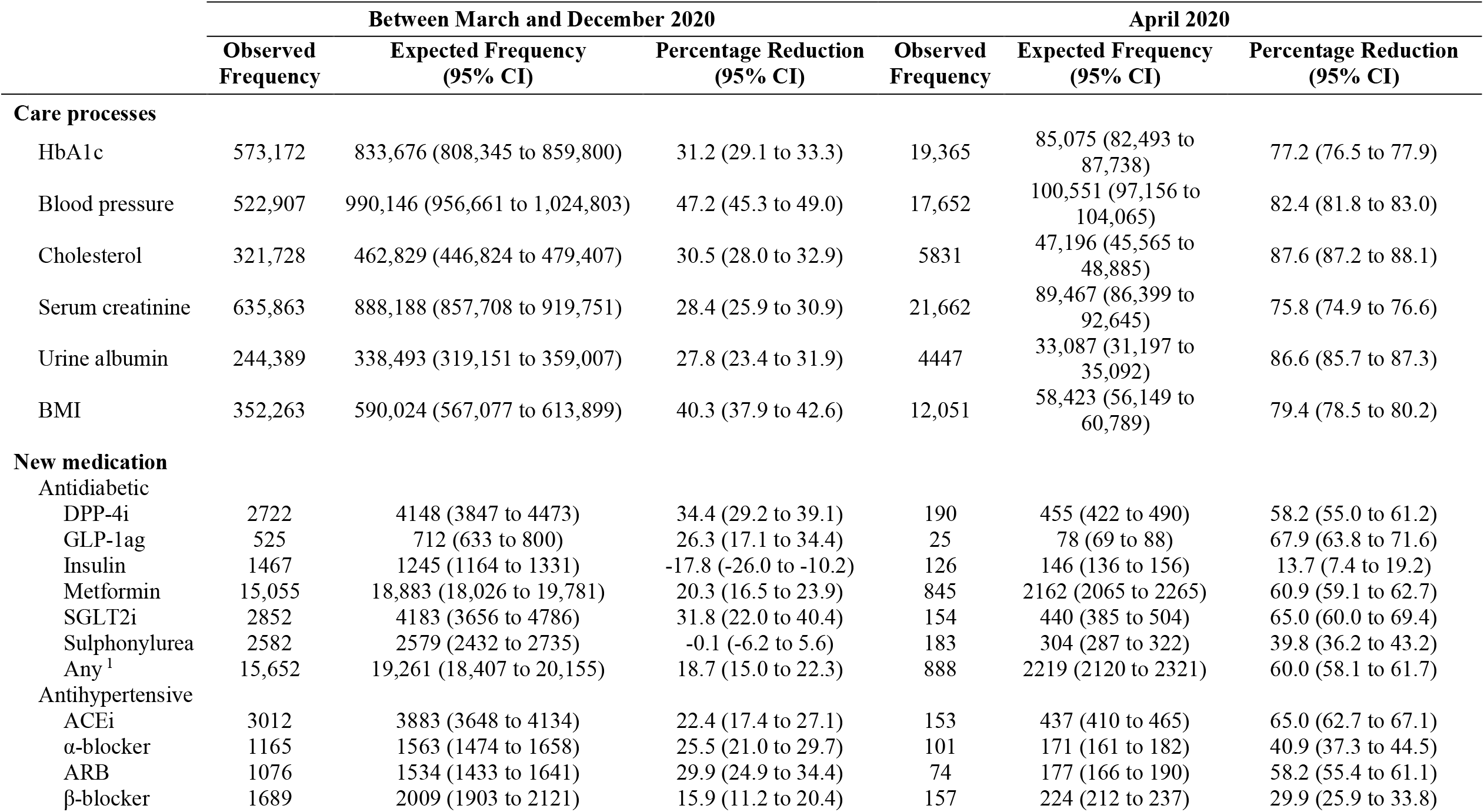
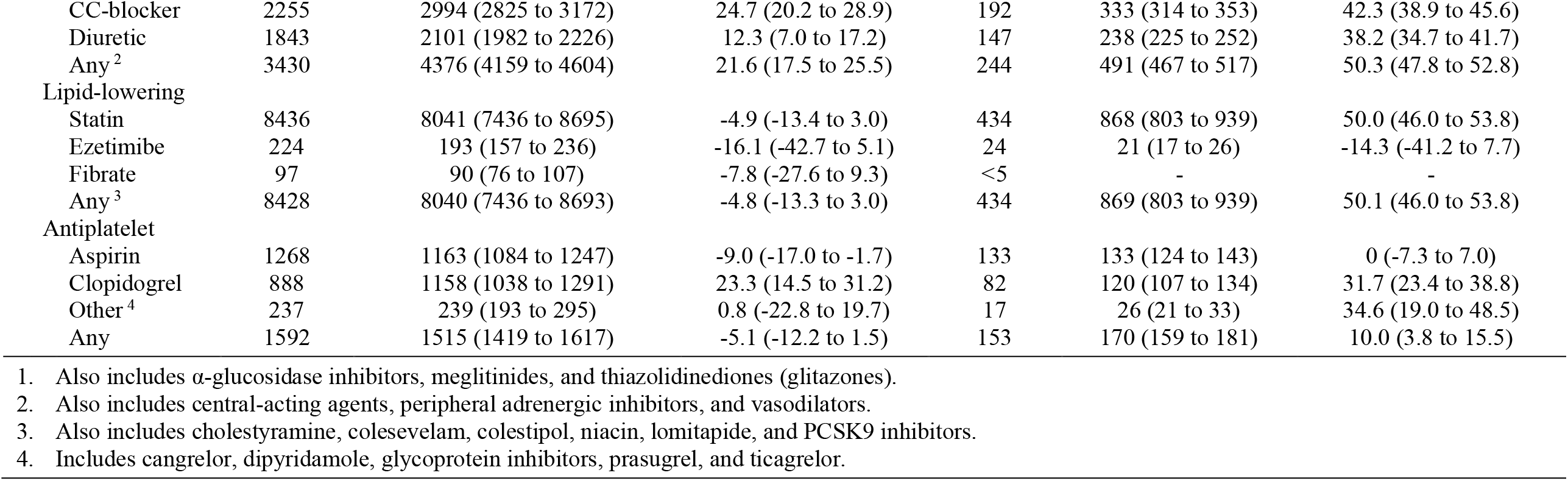
Comparison of observed and expected rates of diabetes-related care process implementation and new medication initiation in people with type 2 diabetes between March and December 2020 and in April 2020, in England

#### Impacts of COVID-19 on diabetes-related prescribing

We assessed changes in rates of prescribing of new diabetes medication along with new antihypertensive, lipid-lowering and antiplatelet medications in people with T2D. In England, prescribing of new medication fell during April with rate reductions varying from 10% (4-16%) for antiplatelet agents to 60% (58-62%) for antidiabetic medications (**Figure 3**). Similar patterns were observed in other UK nations with rate reductions varying between 26% (15-34%) for antiplatelet agents and 64% (61-66%) for lipid-lowering therapy (**Supplementary Figure 3**).

**Figure 3.**
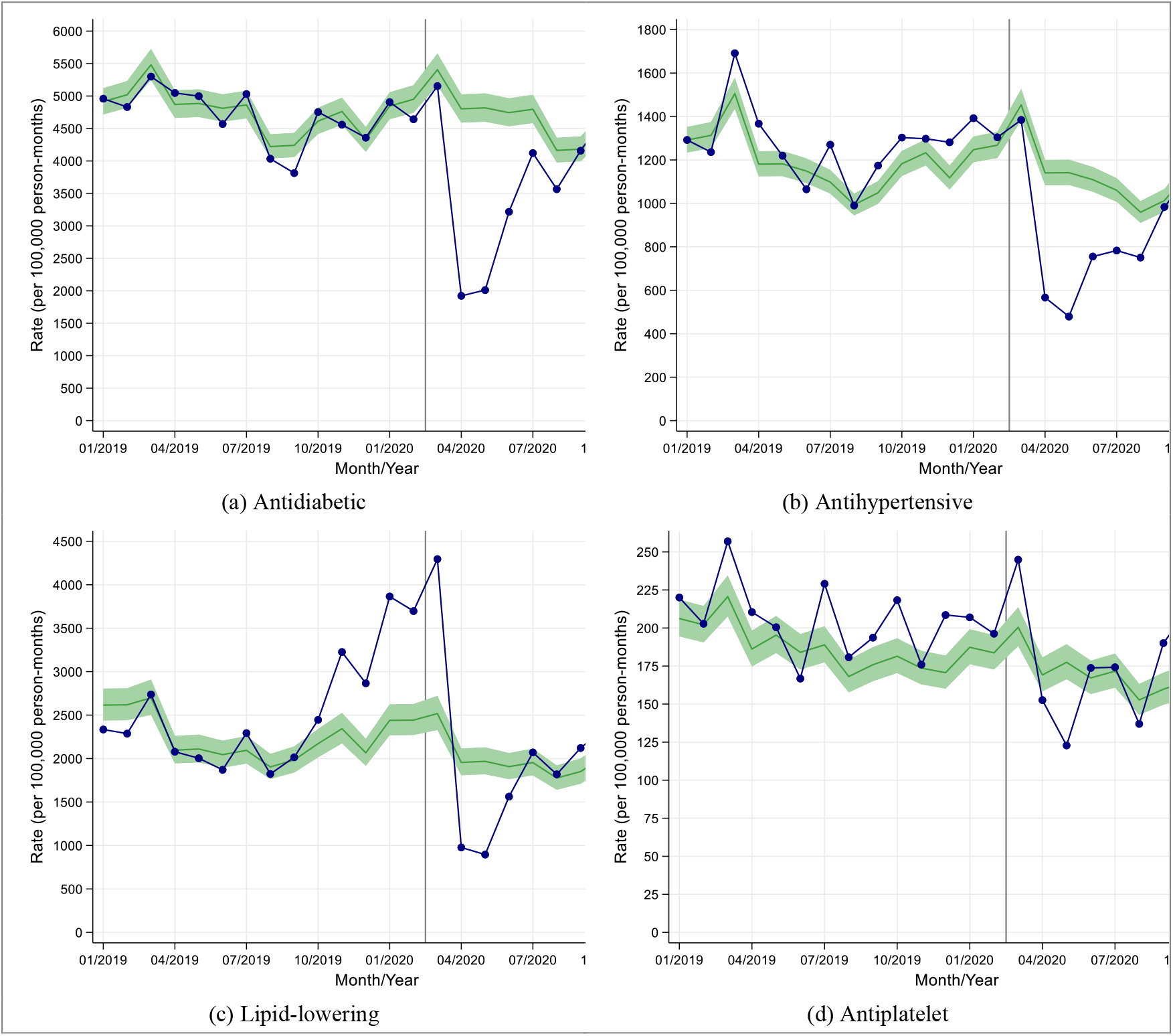
Observed and expected rates of new medication initiation in people with type 2 diabetes during 2019 and 2020, in England

In contrast to the data on rates of performing care processes, the largest reductions in rates of prescribing new diabetes medication and new lipid lowering medication in England were seen in younger individuals from deprived and non-deprived backgrounds (**Supplementary Figure 4**).

Overall in English practices, between 1^st^ March and 10^th^ December 2020, the overall rate of prescribing new diabetes medications was reduced by 19% (15-22%) when compared to historical trends (**Table 1**); the most affected medication being dipeptidyl peptidase-4 inhibitors (reduction: 34% (29-39%)) and the least affected being insulin which was initiated more frequently during this period compared to historical trends (increase: 18% (10-26%)).

Similarly, the prescribing of new antihypertensive medication was reduced by 22% (18-25%) during this period whereas there was no significant change in the prescribing of new lipid lowering or new antiplatelet therapy (**Table 1**).

Between 1^st^ March and 10^th^ December 2020, similar reductions in the trends for prescribing of new medication were observed in other UK nations except that there was a significant reduction in new lipid-lowering therapy prescribing (reduction: 15 (10 to 21)%; **Supplementary Table 2**).

When considering both new prescribing and repeat prescribing combined, there was no significant change in the prescribing of antidiabetic, antihypertensive, lipid-lowering or antiplatelet therapies between March 2020 and December 2020 (England, **Figure 4**; other UK nations, **Supplementary Figure 5**).

**Figure 4.**
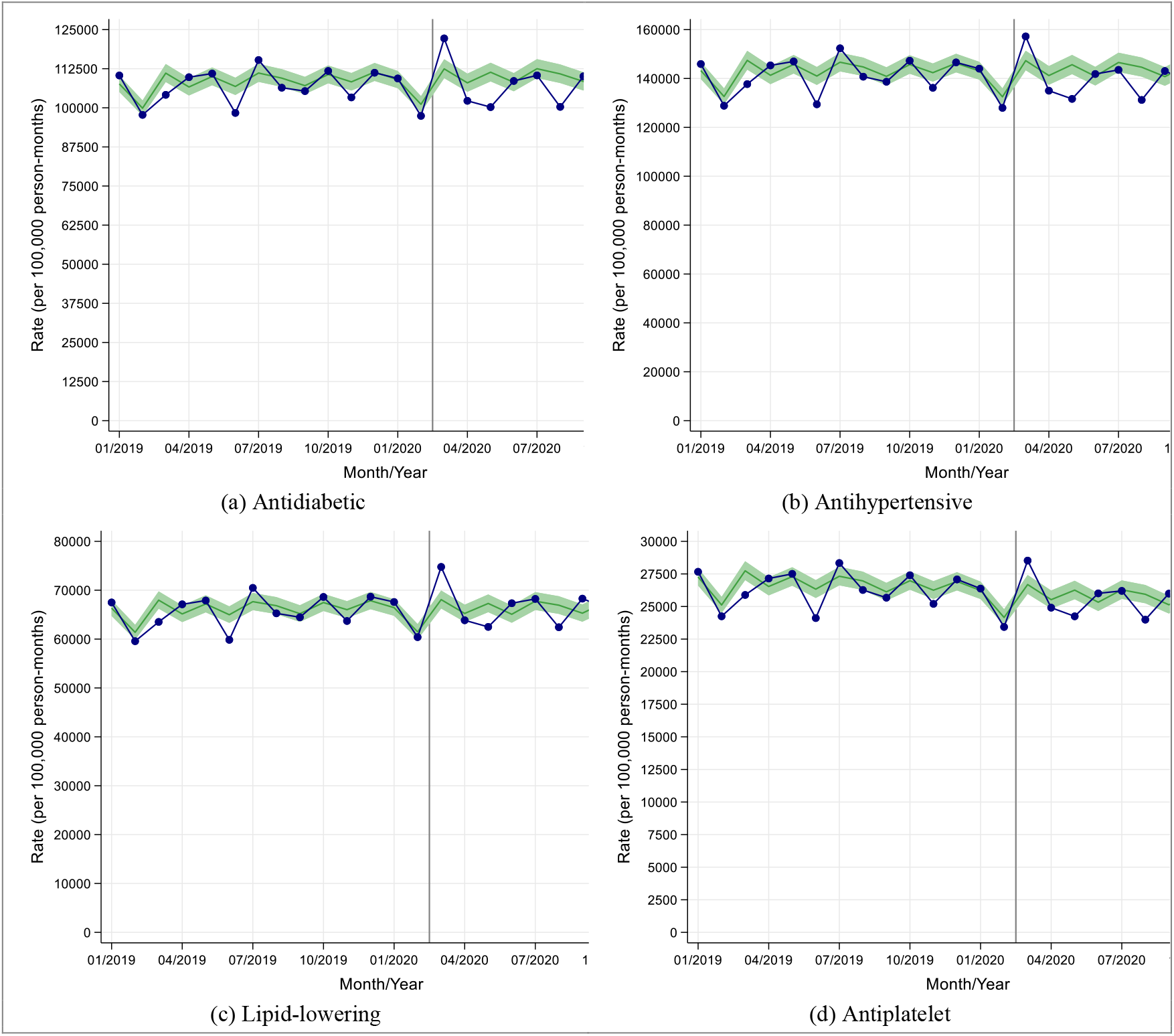
Observed and expected new and repeat medication prescribing rates in people with type 2 diabetes during 2019 and 2020, in England

## DISCUSSION

We used primary care electronic health records from more than 600,000 people with T2D in the UK, and 10-year historical data, to show that during the nine months following the first nationwide ‘lockdown’, the indirect consequences of the COVID-19 pandemic led to clinically significant changes in care quality and prescribing that could adversely impact patient safety. When compared to historical trends, we showed that: i) there were 28-47% reductions in rates of performing a range of health checks including a near halving of blood pressure testing rates; ii) older people with T2D and those from more deprived backgrounds experienced the greatest reduction in health checks; iii) overall rates of prescribing new diabetes and antihypertensive medication were reduced by 19-22%; and iv) reassuringly, when considering rates of new and repeat medication prescribing combined, there were no significant differences.

There are limited data on the impact of the COVID-19 pandemic on rates of performing health checks and prescribing in people with type 2 diabetes. An earlier UK-wide study using primary care data from people with T2D, showed a 31% reduction in HbA1c testing, a 20% reduction in new metformin prescribing and 5% reduction in new insulin prescribing between March and December 2020 compared to historical trends.^7^ Here we extend these observations by assessing the impact of the COVID-19 pandemic on a much wider range of health checks and a wider range of diabetes-related medication including agents that reduce cardiovascular risk.

Our findings have important clinical implications for diabetes care quality and safety. In early March 2020, GPs were advised to minimise the number of face-to-face contacts they had with their patients.^8^ Our data suggests that this reduction of clinical services has led to major reductions in the monitoring of T2D and the prescribing of new medication, particularly for hypertension. T2D is a progressive condition and therefore without intervention, levels of glucose and associated CVD risk factors such as blood pressure tend to increase over time. There are already concerns about clinical inertia in diabetes management in the UK,^9^ and therefore any further reduction in monitoring and related prescribing could increase the risk of mortality and long-term complications.

Our data indicate that reductions in prescribing relate to new prescriptions but not repeat prescriptions. Robust systems for repeat prescribing in the UK appear to have helped minimise the harm done through reductions in face-to-face consultations during the pandemic.

In addition to the reduction in care quality and the potential risks posed by reduced monitoring and prescribing, there is evidence that national lockdowns have had additional detrimental effects on cardiovascular disease (CVD) risk especially in people with T2D. In surveys of UK adults conducted during the first lockdown (April-May 2020), participants reported adverse changes in several behaviours that promote weight gain (adverse changes in diet, physical activity, alcohol consumption, mental health and sleep quality).^10 11^ Other studies in people with T2D have shown worsening of glycaemic control in relation to COVID-19 lockdown.^12 13^

As engagement with health services increases over the coming months, we predict marked increases in the need for monitoring and prescribing of new medication in people with T2D. Healthcare services will need to manage this backlog of testing and prescribing, and the anticipated greater deterioration of HbA1c and other CVD risk factors such as blood pressure levels. Older people from deprived backgrounds appear to be most adversely affected by reduced monitoring and therefore these individuals may be particular groups to target for early intervention. During this pandemic and its associated lockdowns, effective communications should ensure that patients remain engaged with diabetes services, monitoring and opportunities for prescribing,^14^ and make use of home monitoring of blood pressure, weight and HbA1c (when available), and remote consultations.^15-17^ Investment in such technology and devices would be expected to yield important health benefits.

The COIVD-19 pandemic provides us with a unique opportunity to improve the current care model by providing greater investment in patient education, devices and technology and greater use of remote consultations to deliver the high standards of care that people with diabetes should expect to receive.

Our study had several strengths: this is the first UK-wide study reporting the indirect impact of the COVID-19 pandemic on health checks and prescribing in people with T2D. Our findings in English practices were replicated in other parts of the UK and are likely to be representative of the UK in general. Our study has some limitations: First, we did not report data on retinopathy, smoking and foot checks (the remaining 3 of the 9 health checks recommended by NICE. Retinopathy screening is performed in the community and therefore these data are not available in primary care records. Whist assessments of smoking status and foot checks are performed in primary care, we were less confident in defining whether or not these checks had been performed based on the available data. Second, we do not present data on type 1 diabetes as the majority of care for these individuals is delivered in secondary care centres. Third, ethnicity coding is not adequately captured in primary care and therefore we had limited ability to explore ethnicity-related variation in outcomes. Fourth, we did not assess risk factor levels because our focus was on processes of care and prescribing. Fifth, our data would not capture assessments of weight and blood pressure assessed by patients in their homes. Results of home blood pressure recordings may have had an influence on prescribing between March and December 2020 because the reduction in prescribing of new antihypertensive agents (∼13%) was less than the reduction in BP monitoring performed in primary care (∼51%). Finally, although our results and conclusions are relevant to the UK population, generalisability to other healthcare systems may be limited. However, a pan-European survey of diabetes specialist nurses reported that the level of care provided for people with diabetes had been significantly disrupted during the pandemic.^25^

In conclusion, we highlight marked reductions in the rate of health checks and new prescribing in people with T2D as indirect consequences of the COVID-19 pandemic. Over the coming months, healthcare services will need to manage this backlog of testing and prescribing, and the anticipated greater deterioration of HbA1c and other CVD risk factors such as blood pressure levels. Older people from deprived backgrounds with T2D may be specific groups to target for early health checks and intervention. During the remainder of this pandemic, effective communications should ensure that patients remain engaged with diabetes services, monitoring and opportunities for prescribing, and make use of home blood pressure, weight and HbA1c monitoring when available. Healthcare planners should seize opportunities provided by the COVID-19 pandemic to improve models, processes and standards of care for people with diabetes.

## Data Availability

All clinical codes used in the study are published on Clinicalcodes.org. Electronic health records are, by definition, considered sensitive data in the UK by the Data Protection Act and cannot be shared via public deposition because of information governance restriction in place to protect patient confidentiality. Access to data are available only once approval has been obtained through the individual constituent entities controlling access to the data. The primary care data can be requested via application to the Clinical Practice Research Datalink (www.cprd.com).

## Acknowledgements

The study is based on data from the Clinical Practice Research Datalink obtained under license from the UK Medicines and Healthcare products Regulatory Agency. The study and use of CPRD data was approved by the Independent Scientific Advisory Committee for Clinical Practice Research Datalink research (protocol number: 20_182R). The data is provided by patients and collected by the NHS as part of their care and support. The interpretation and conclusions contained in this study are those of the authors alone. We would like to acknowledge all the data providers and general practices that made the anonymised data available for research.

## Author Contributions

DMA conceived the original idea. MKR, MJC and AKW helped develop the idea. MJC and AKW performed the analysis and verified the analytical methods. MKR and DMA also reviewed the clinical code sets. NK and NM (primary care clinicians) and LL, HT (secondary care diabetes clinicians) helped interpret the results. MKR wrote the manuscript with input from all authors. All authors critically reviewed and approved the final version.

The lead authors (MJC and AKW: the manuscript’s guarantors) affirm that the manuscript is an honest, accurate, and transparent account of the study being reported; that no important aspects of the study have been omitted; and that any discrepancies from the study as planned have been explained. The corresponding author had full access to all of the data and the final responsibility to submit for publication.

## Data sharing

All clinical codes used in the study are published on Clinicalcodes.org. Electronic health records are, by definition, considered “sensitive” data in the UK by the Data Protection Act and cannot be shared via public deposition because of information governance restriction in place to protect patient confidentiality. Access to data are available only once approval has been obtained through the individual constituent entities controlling access to the data. The primary care data can be requested via application to the Clinical Practice Research Datalink (www.cprd.com).

