## Supplementary for "Impact of COVID-19 restrictions on diabetes health checks and prescribing for people with type 2 diabetes: a UK-wide cohort study involving 618,161 people in primary care"

### Supplemental Material

**Table of Contents**

**Supplementary Figure 1.** Observed and expected care process rates in people with type 2 diabetes during 2019 and 2020, in Northern Ireland, Scotland and Wales 2

**Supplementary Figure 2.** Stratified care process rates in people with type 2 diabetes during 2019 and 2020, in Northern Ireland, Scotland and Wales 3

**Supplementary Table 1.** Strata applied to study cohorts 4

**Supplementary Table 2.** Comparison of observed and expected rates of diabetes-related care process implementation and new medication initiation in people with type 2 diabetes between March and December 2020 and in April 2020, in Northern Ireland, Scotland, and Wales 5

**Supplementary Figure 3.** Observed and expected rates of new medication initiation in people with type 2 diabetes during 2019 and 2020, in Northern Ireland, Scotland and Wales 6

**Supplementary Figure 4.** Stratified rates of new medication initiation in people with type 2 diabetes during 2019 and 2020, in England 7

**Supplementary Figure 5.** Observed and expected new and repeat medication prescribing rates in people with type 2 diabetes during 2019 and 2020, in Northern Ireland, Scotland and Wales 8

**Supplementary Figure 1.** Observed and expected care process rates in people with type 2 diabetes during 2019 and 2020, in Northern Ireland, Scotland and Wales

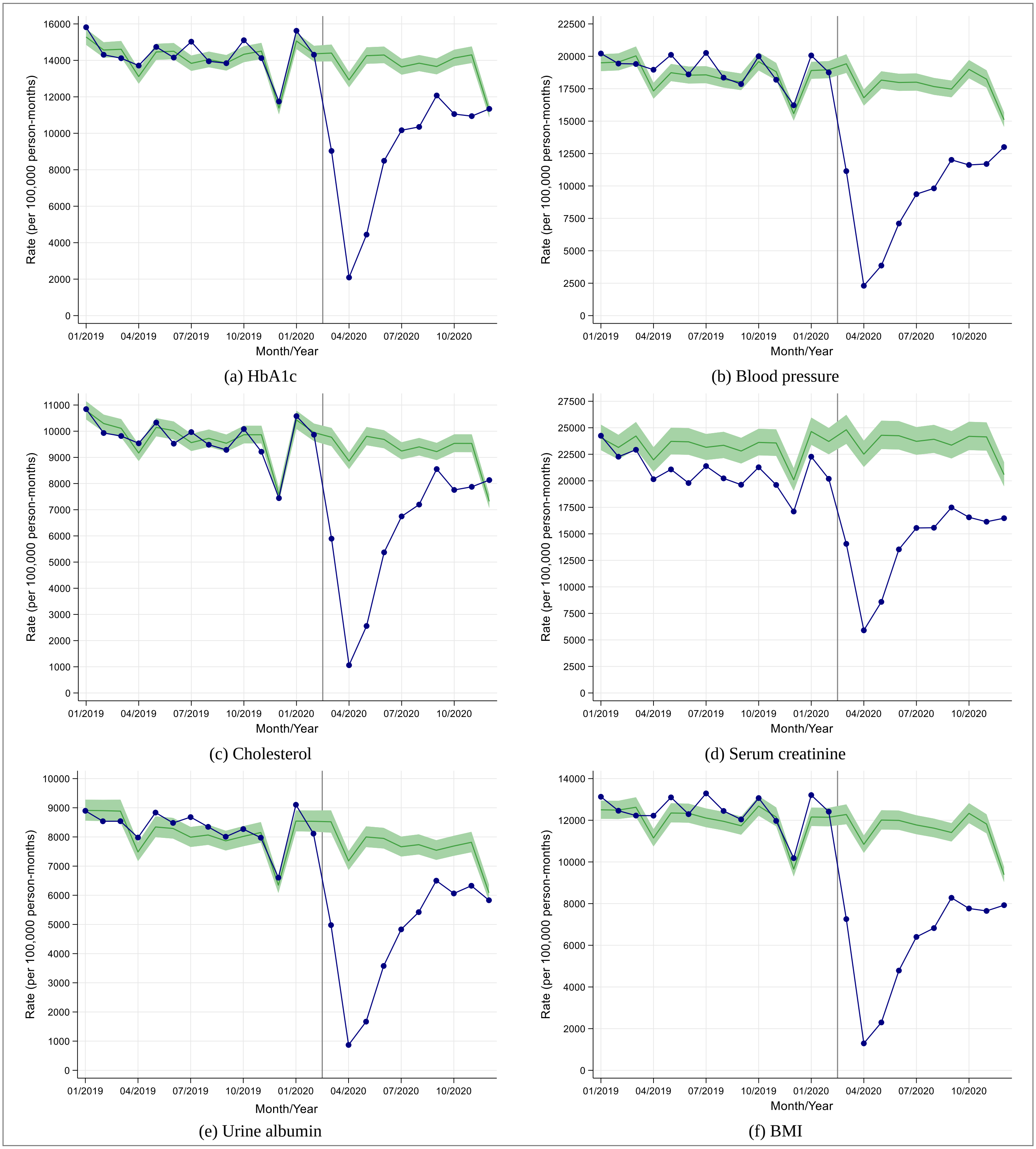


(a) HbA1c

(b) Blood pressure

(c) Cholesterol

(d) Serum creatinine

(e) Urine albumin

(f) BMI

**Supplementary Figure 2.** Stratified care process rates in people with type 2 diabetes during 2019 and 2020, in Northern Ireland, Scotland and Wales

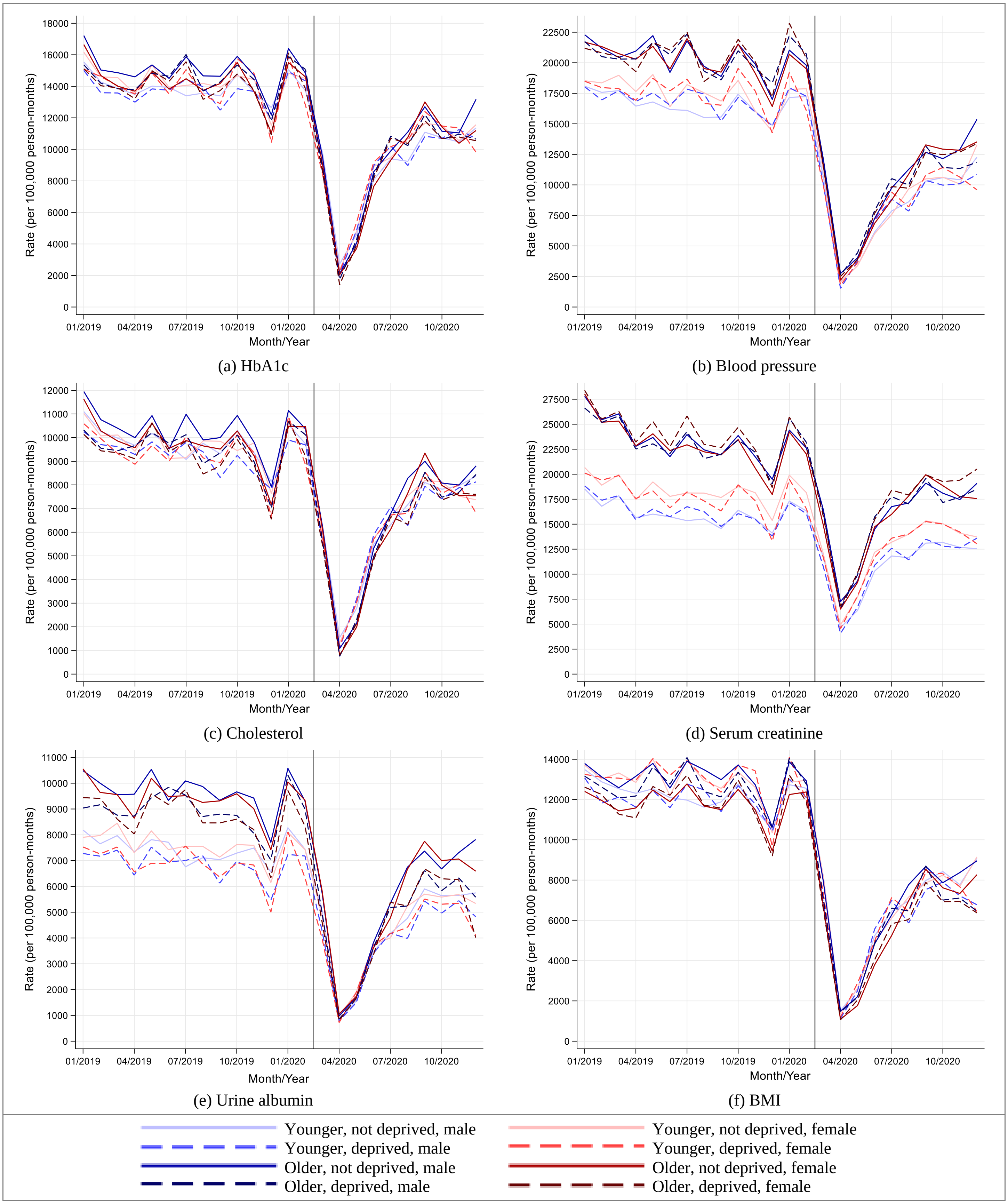

**Supplementary Table 1.** Strata applied to study cohorts

| Stratum | Label | Older | Deprived | Female |
|---|---|---|---|---|
| 1 | Younger, not deprived, male | 0 | 0 | 0 |
| 2 | Younger, not deprived, female | 0 | 0 | 1 |
| 3 | Younger, deprived male | 0 | 1 | 0 |
| 4 | Younger, deprived female | 0 | 1 | 1 |
| 5 | Older, not deprived, male | 1 | 0 | 0 |
| 6 | Older, not deprived, female | 1 | 0 | 1 |
| 7 | Older, deprived male | 1 | 1 | 0 |
| 8 | Older, deprived female | 1 | 1 | 1 |

- Older = 0,1 if age ≥ 65
- Deprived = 0,1 if IMD quintile = 4 or 5
- Female = 0,1

**Supplementary Table 2.** Comparison of observed and expected rates of diabetes-related care process implementation and new medication initiation in people with type 2 diabetes between March and December 2020 and in April 2020, in Northern Ireland, Scotland, and Wales

| | Between March and December 2020 | | | April 2020 | | |
|---|---|---|---|---|---|---|
| | **Observed Frequency** | **Expected Frequency (95% CI)** | **Percentage Reduction (95% CI)** | **Observed Frequency** | **Expected Frequency (95% CI)** | **Percentage Reduction (95% CI)** |
| **Care processes** | | | | | | |
| HbA1c | 72,084 | 113,705 (110,149 to 117,376) | 36.6 (34.6 to 38.6) | 1862 | 11,513 (11,153 to 11,885) | 83.8 (83.3 to 84.3) |
| Blood pressure | 72,650 | 147,623 (142,287 to 153,159) | 50.8 (48.9 to 52.6) | 2050 | 14,967 (14,427 to 15,526) | 86.3 (85.8 to 86.8) |
| Cholesterol | 48,674 | 77,035 (74,341 to 79,826) | 36.8 (34.5 to 39.0) | 944 | 7891 (7615 to 8177) | 88.0 (87.6 to 88.5) |
| Serum creatinine | 112,867 | 195,299 (184,827 to 206,364) | 42.2 (38.9 to 45.3) | 5260 | 20,049 (18,976 to 21,182) | 73.8 (72.3 to 75.2) |
| Urine albumin | 36,881 | 63,500 (60,730 to 66,396) | 41.9 (39.3 to 44.5) | 774 | 6393 (6115 to 6684) | 87.9 (87.3 to 88.4) |
| BMI | 48,242 | 96,139 (92,499 to 99,922) | 49.8 (47.8 to 51.7) | 1151 | 9659 (9293 to 10,038) | 88.1 (87.6 to 88.5) |
| **New medication** | | | | | | |
| Antidiabetic | | | | | | |
| DPP-4i | 414 | 619 (566 to 677) | 33.1 (26.9 to 38.8) | 27 | 68 (62 to 74) | 60.3 (56.5 to 63.5) |
| GLP-1ag | 85 | 82 (67 to 102) | -3.7 (-26.9 to 16.7) | <5 | - | - |
| Insulin | 234 | 224 (195 to 258) | -4.5 (-20.0 to 9.3) | 21 | 23 (20 to 27) | 8.7 (-5.0 to 22.2) |
| Metformin | 2874 | 3310 (3121 to 3510) | 13.2 (7.9 to 18.1) | 138 | 386 (364 to 409) | 64.2 (62.1 to 66.3) |
| SGLT2i | 616 | 929 (806 to 1070) | 33.7 (23.6 to 42.4) | 26 | 92 (79 to 107) | 71.7 (67.1 to 75.7) |
| Sulphonylurea | 599 | 625 (578 to 675) | 4.2 (-3.6 to 11.3) | 41 | 75 (69 to 81) | 45.3 (40.6 to 49.4) |
| Any [1] | 2985 | 3417 (3224 to 3622) | 12.6 (7.4 to 17.6) | 147 | 403 (381 to 427) | 63.5 (61.4 to 65.6) |
| Antihypertensive | | | | | | |
| ACEi | 526 | 685 (636 to 737) | 23.2 (17.3 to 28.6) | 23 | 77 (71 to 83) | 70.1 (67.6 to 72.3) |
| α-blocker | 214 | 249 (220 to 282) | 14.1 (2.7 to 24.1) | 11 | 26 (22 to 29) | 57.7 (50.0 to 62.1) |
| ARB | 180 | 223 (194 to 256) | 19.3 (7.2 to 29.7) | 9 | 24 (21 to 28) | 62.5 (57.1 to 67.9) |
| β-blocker | 328 | 384 (342 to 432) | 14.6 (4.1 to 24.1) | 21 | 39 (34 to 44) | 46.2 (38.2 to 52.3) |
| CC-blocker | 363 | 472 (430 to 518) | 23.1 (15.6 to 29.9) | 22 | 56 (51 to 61) | 60.7 (56.9 to 63.9) |
| Diuretic | 375 | 384 (344 to 428) | 2.3 (-9.0 to 12.4) | 20 | 42 (37 to 46) | 52.4 (45.9 to 56.5) |
| Any [2] | 616 | 738 (684 to 795) | 16.5 (9.9 to 22.5) | 40 | 81 (75 to 87) | 50.6 (46.7 to 54.0) |
| Lipid-lowering | | | | | | |
| Statin | 1196 | 1420 (1325 to 1522) | 15.8 (9.7 to 21.4) | 55 | 153 (143 to 165) | 64.1 (61.5 to 66.7) |
| Ezetimibe | 33 | 18 (11 to 29) | -83.3 (-200.0 to -13.8) | <5 | - | - |
| Fibrate | 25 | 16 (11 to 23) | -56.3 (-127.3 to -8.7) | <5 | - | - |
| Any [3] | 1199 | 1419 (1325 to 1520) | 15.5 (9.5 to 21.1) | 56 | 153 (143 to 164) | 63.4 (60.8 to 65.9) |
| Antiplatelet | | | | | | |
| Aspirin | 239 | 229 (202 to 260) | -4.4 (-18.3 to 8.1) | 16 | 24 (21 to 27) | 33.3 (23.8 to 40.7) |
| Clopidogrel | 196 | 228 (192 to 271) | 14.0 (-2.1 to 27.7) | 14 | 23 (19 to 27) | 39.1 (26.3 to 48.1) |
| Other [4] | 61 | 61 (42 to 88) | 0 (-45.2 to 30.7) | 5 | 5 (4 to 8) | 0 (-25.0 to 37.5) |
| Any | 300 | 298 (266 to 336) | -0.7 (-12.8 to 10.7) | 23 | 31 (27 to 35) | 25.8 (14.8 to 34.3) |

1. Also includes α-glucosidase inhibitors, meglitinides, and thiazolidinediones (glitazones).
2. Also includes central-acting agents, peripheral adrenergic inhibitors, and vasodilators.
3. Also includes cholestyramine, colesevelam, colestipol, niacin, lomitapide, and PCSK9 inhibitors.
4. Includes cangrelor, dipyridamole, glycoprotein inhibitors, prasugrel, and ticagrelor.

**Supplementary Figure 3.** Observed and expected rates of new medication initiation in people with type 2 diabetes during 2019 and 2020, in Northern Ireland, Scotland and Wales

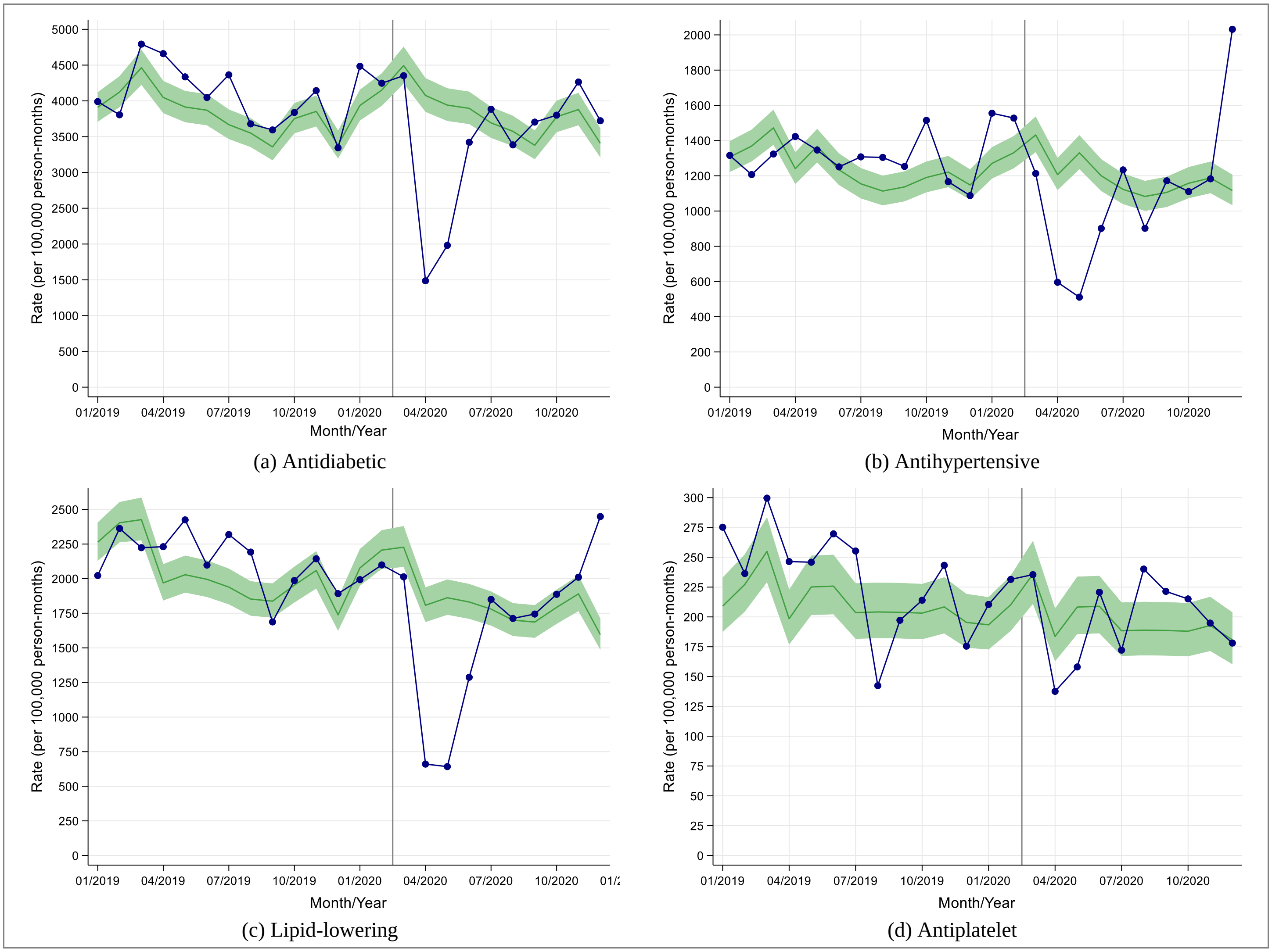


(a) Antidiabetic

(b) Antihypertensive

(c) Lipid-lowering

(d) Antiplatelet

**Supplementary Figure 4.** Stratified rates of new medication initiation in people with type 2 diabetes during 2019 and 2020, in England

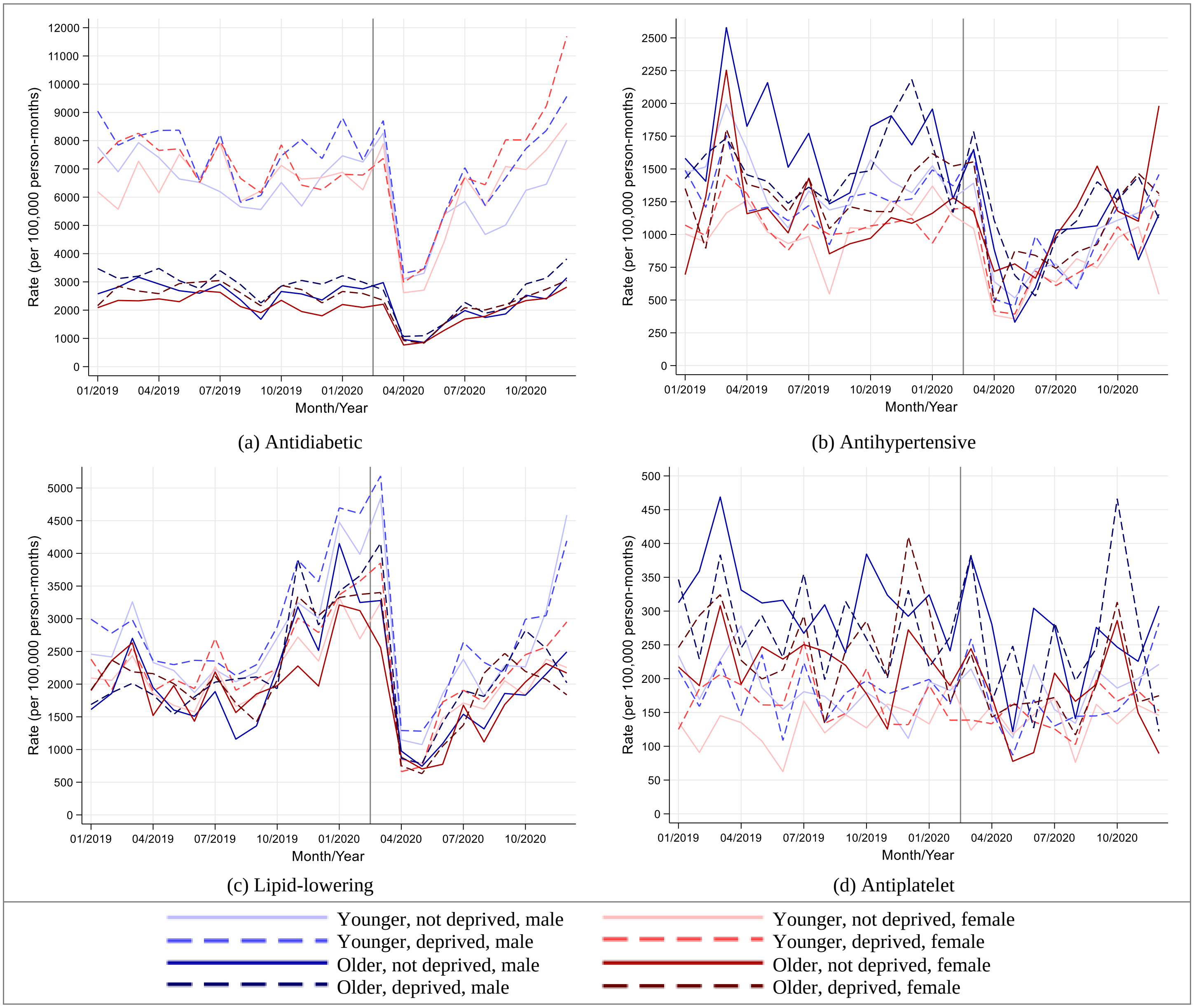

**Supplementary Figure 5.** Observed and expected new and repeat medication prescribing rates in people with type 2 diabetes during 2019 and 2020, in Northern Ireland, Scotland and Wales

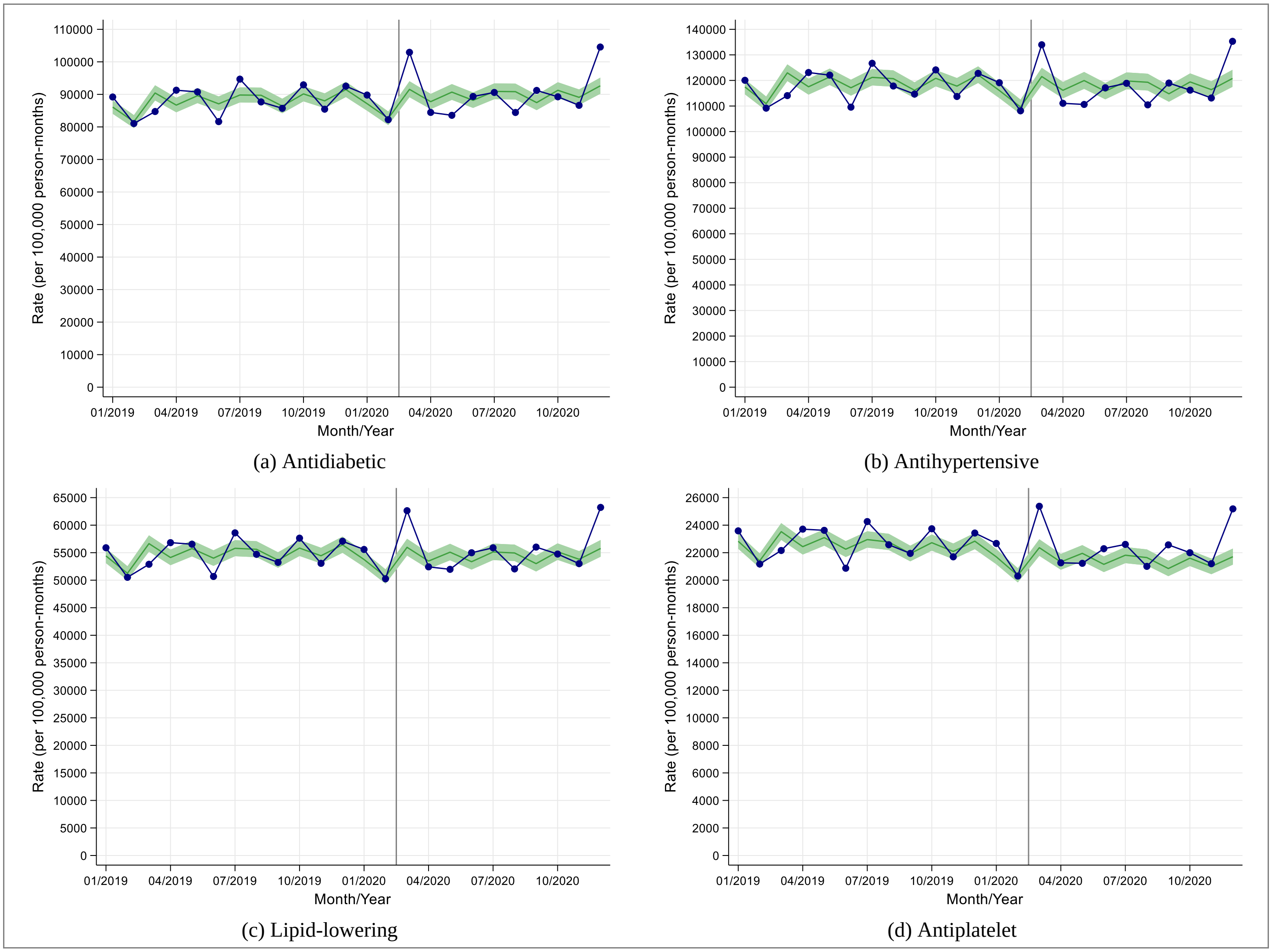


(a) Antidiabetic

(b) Antihypertensive

(c) Lipid-lowering

(d) Antiplatelet
